# Cardiometabolic Indicators of Cognitive Impairment in The Cameron County Hispanic Cohort

**DOI:** 10.1101/2025.01.08.25320180

**Authors:** Fadi I. Musfee, Seema Agarwal, Vahed Maroufy, Joseph McCormick, Susan Fisher-Hoch, Sean I. Savitz

## Abstract

**Introduction:** Cognitive impairment (CI) and its related risk factors (e.g., diabetes and stroke) are highly prevalent among Hispanic/Latinos (H/L); however, prior research in H/L focused on aging individuals (≥65 years of age). We conducted a prospective study in a younger cohort of H/L (majority <65 years old) from the Cameron County Hispanic Cohort (CCHC) to comprehensively assess the associations between a wide-range of cardiometabolic health indicators with CI.

**Methods:** We identified a total of 1240 CCHC subjects with complete Mini-mental status exam (MMSE) data at study baseline and at 5-year follow-up. The outcome (i.e., CI) was based on MMSE scores of less than 24. We conducted univariate associations for multiple cardiometabolic indicators with CI; and mixed logistic regression models to estimate odds ratios for the associations between cardiometabolic indicators and CI adjusted for age, education, prior stroke, and *APOE* gene.

**Results:** The majority (89.9%) of the participants were <65 years old. A total of 117 subjects had CI at baseline (9.4%). Baseline study cohort showed that Individuals with CI were older with a lower education performance, and were more likely to be diabetic with lower mean levels of Low-density Lipoprotein, and a higher mean systolic blood pressure. Diabetes significantly increased the odds for CI (OR:2.11, 95%CI:1.26-3.52) from the adjusted multivariate mixed logistic models.

**Conclusions:** This analysis showed that diabetes was an important indicator for CI regardless of age, education, or *APOE* gene status. These findings highlight the higher burden of cardiometabolic risk factors on CI in the CCHC cohort.

## Introduction

Dementia is a collective term that describes a steady decline in the individual cognitive abilities with a prevalence of over 55 million individuals worldwide [1]. The course of dementia usually starts with a transitioning phase of mild cognitive impairment (MCI) that deteriorates over time [1]. Cognitive impairment that develops in the presence of vascular risk factors such as hypertension and diabetes, or as a result of cerebrovascular disease due to ischemic or hemorrhagic stroke is referred to as vascular cognitive impairment [2].

Hispanics/Latinos (H/L) tend to have a higher incidence of dementia and MCI when compared to Non-Hispanic (NH) Whites [2]. Similarly, H/L tend to have a higher incidence of vascular risk factors compared to NH Whites which influence the development of vascular dementia and vascular MCI [2]. Furthermore, H/L tend to have a different genetic background compared to NH white populations. In particular, H/L populations showed weaker *APOE* gene associations with dementia, as far less cases of dementia among H/L were explained by the *APOE* haplotype compared to NH Whites [3–5]. Additionally, genetic risk scores for diabetes have not been associated with increased risk for dementia in H/L, as was the case with NH White [6]. These differences in genetic backgrounds make the case for a higher influence of cardiometabolic risk factors as drivers for the high incidence of cognitive impairment in H/L [7]. Lastly, H/L, mainly Mexican Americans, tend to develop cognitive impairment at a younger age group when compared to NH Whites [5]. Prior research studies in dementia in the H/L population have focused on those over 65 years of age and mainly addressed the relationship between dementia and other cardiovascular risk factors in cross-sectional or case-control study designs [7].

The objective of this study was to investigate the associations of multiple demographics and cardiometabolic indicators with incident mild cognitive impairment in the Cameron County Hispanic Cohort (CCHC), a longitudinal cohort study of individuals living in Cameron County in Brownsville, Texas mainly consisting of Mexican origin H/L who are selected randomly to participate in this cohort. We hypothesize that cardiometabolic factors are the major drivers for the high incidence of cognitive impairment in H/L populations of Mexican origin and the CCHC in particular.

## Methods

The CCHC, a longitudinal study that collects data from the local H/L community in the Rio Grande Valley, is mainly consisted of Mexican American participants. Details regarding the CCHC’s randomization, recruitment, and data collection procedures have been described previously [8]. Participants were recruited based on weighted sampling to achieve a representative sample of the local community. Our longitudinal study analyses were based on a subset of the participants in the CCHC cohort who received a Mini-Mental State Examination (MMSE) assessment at the baseline interview and a follow up MMSE assessment at their second visit (i.e., 5-year follow-up interview).

Consented CCHC participants provided their medical and socioeconomic information through standardized questionnaires which included the MMSE assessment at baseline and each following visit. Participants answered those questionnaires in either English or Spanish, based on their preferred language. Blood samples were also collected at the baseline interview and at each following visit. This study was approved by the Institutional Review Board (IRB) of the University of Texas Health Science Center at Houston.

### Sample selection

Currently, there are more than 5,000 CCHC participants with data points of follow-up for 20 years (2004 – 2024). To construct our prospective cohort, we limited the current analysis to include those who had their second follow up visit after 5 years from their baseline enrollment (i.e., each patient in this cohort will have two visits: a baseline visit, and a second visit after 5 years). The outcome (i.e., cognitive impairment) of this study was defined based on MMSE score where MMSE of less than 24 was considered as an indication of cognitive impairment [9]. This MMSE score was determined based on clinical trials practices which applied MMSE scores to select participants with Alzheimer disease [9]. Clinical trials excluded participants with cognitive impairment from receiving disease modifying treatment for Alzheimer disease using MMSE scores of a mean of 24 points [9]. Additionally, subjects were excluded from our cohort based on age (<18 years of age were excluded), and the presence of MMSE scores data (subjects with missing MMSE scores were excluded).

### Statistical Analysis

All analyses were conducted using R software (version 4.4.2). We included independent variables and risk factors that were tested in prior research studies on cognition and Alzheimer disease from Hispanic and Non-Hispanic populations. We conducted univariate associations at study baseline for multiple cardiometabolic indicators (e.g., hypertension [systolic and diastolic], diabetes [diabetes history and HbA1c], lipids [Low Density Lipoproteins (LDL-c), High Density Lipoproteins (HDL-c), Triglyceride, Total Cholesterol], echocardiogram measurements [Left Ventricular Ejection fraction (LVEF), Left Ventricular End Diastolic Diameter (LVIDd), Left Ventricular End Systolic Diameter (LVIDs), and Left Ventricular (LV) mass], inflammatory markers [C reactive protein (CRP), IL1beta, IL6, IL8, Tumor Necrosis Factor (TNF) alpha), and sociodemographic factors (e.g., age, education level, Body mass index, physical activity, annual income) with cognitive impairment using Chi square, Fisher exact, and student T-test. Next, we ran mixed logistic regression models to estimate the odds ratios for the associations between several cardiometabolic indicators and cognitive impairment adjusted for age, education, prior stroke, and *APOE* haplotypes. To construct the 95% confidence intervals and p values that accounts for the random effect from the mixed logistic model, we estimated the random standard error using the variance-covariance matrix from the mixed logistic regression analysis. The definitions and criteria used in reporting those risk factors in the CCHC cohort are provided previously [9]. We excluded risk factors with more than 25% of missing information from our mixed logistic models. Table 1. To identify the level of significance with multiple testing, we used False discovery rate (FDR) method to estimate the threshold of significance for multiple testing (p value<0.05). Finally, we looked into the dose-response relationship between Hemoglobin A1c and MMSE scores using adjusted and unadjusted mixed logistic modeling.

**Table 1.** Cohort characteristics showing the distributions for multiple cardiometabolic health indicators at study baseline.

| Risk Factor | Normal Cognition<br>(N=1123) | Cognitive<br>Impairment<br>(N=117) | Overall<br>(N=1240) | p value |
| --- | --- | --- | --- | --- |
| **Plaque presence on Carotid Intima-Media Thickness** |  |  |  |  |
| Yes | 42 (3.7%) | 4 (3.4%) | 46 (3.7%) | 0.72 |
| No | 218 (19.4%) | 14 (12.0%) | 232 (18.7%) |  |
| Missing | 863 (76.8%) | 99 (84.6%) | 962 (77.6%) |  |
| **Flow mediated dilation (FMD)** |  |  |  |  |
| Abnormal | 71 (6.3%) | 6 (5.1%) | 77 (6.2%) | 0.99 |
| Normal | 69 (6.1%) | 6 (5.1%) | 75 (6.0%) |  |
| Missing | 983 (87.5%) | 105 (89.7%) | 1088 (87.7%) |  |
| **Age at Visit** |  |  |  |  |
| 18-49 | 663 (59.0%) | 38 (32.5%) | 701 (56.5%) | <0.0001 |
| 50-64 | 364 (32.4%) | 50 (42.7%) | 414 (33.4%) |  |
| >=65 | 96 (8.5%) | 29 (24.8%) | 125 (10.1%) |  |
| **History of Heart Attack** |  |  |  |  |
| Yes | 17 (1.5%) | 4 (3.4%) | 21 (1.7%) | 0.25 |
| No | 1106 (98.5%) | 113 (96.6%) | 1219 (98.3%) |  |
| **History of Stroke** |  |  |  |  |
| Yes | 17 (1.5%) | 7 (6.0%) | 24 (1.9%) | 0.03 |
| No | 1105 (98.4%) | 110 (94.0%) | 1215 (98.0%) |  |
| Missing | 1 (0.1%) | 0 (0%) | 1 (0.1%) |  |
| **Current Alcohol Consumption** |  |  |  |  |
| No | 571 (50.8%) | 70 (59.8%) | 641 (51.7%) | 0.08 |
| Yes | 551 (49.0%) | 47 (40.2%) | 598 (48.2%) |  |
| Missing | 1 (0.1%) | 0 (0%) | 1 (0.1%) |  |
| **Sex** |  |  |  |  |
| Male | 362 (32.2%) | 37 (31.6%) | 399 (32.2%) | 0.97 |
| Female | 761 (67.8%) | 80 (68.4%) | 841 (67.8%) |  |
| **Years of Education (YOE)** |  |  |  |  |
| ≥13 YOE | 349 (31.1%) | 6 (5.1%) | 355 (28.6%) | <0.0001 |
| 9-12 YOE | 446 (39.7%) | 18 (15.4%) | 464 (37.4%) |  |
| 5-8 YOE | 277 (24.7%) | 41 (35.0%) | 318 (25.6%) |  |
| 0-4 YOE | 50 (4.5%) | 52 (44.4%) | 102 (8.2%) |  |
| Missing | 1 (0.1%) | 0 (0%) | 1 (0.1%) |  |
| **Diabetes based on HBA1c categories** |  |  |  |  |
| 1: A1c_NORMAL (GHB < 5.7) | 624 (55.6%) | 52 (44.4%) | 676 (54.5%) | 0.0003 |
| 2: A1c_IMPAIRED (5.7 <= GHB < 6.5) | 282 (25.1%) | 24 (20.5%) | 306 (24.7%) |  |
| 3: A1c_DIABETIC (GHB >=6.5) | 195 (17.4%) | 38 (32.5%) | 233 (18.8%) |  |
| Missing | 22 (2.0%) | 3 (2.6%) | 25 (2.0%) |  |
| **Diabetes (self-reported DM or Taking Medicines for DM or HBA1c >=6.5)** |  |  |  |  |
| No | 868 (77.3%) | 65 (55.6%) | 933 (75.2%) | <0.0001 |
| Yes | 238 (21.2%) | 50 (42.7%) | 288 (23.2%) |  |
| Missing | 17 (1.5%) | 2 (1.7%) | 19 (1.5%) |  |
| **High Blood Pressure (self-reported diagnosis and/or taking medications)** |  |  |  |  |
| Yes | 267 (23.8%) | 50 (42.7%) | 317 (25.6%) |  |
| No | 856 (76.2%) | 67 (57.3%) | 923 (74.4%) |  |
| **Body mass index (BMI categorical)** |  |  |  |  |
| 1: Underweight (BMI < 18.5) | 7 (0.6%) | 0 (0%) | 7 (0.6%) | 0.05 |
| 2: Normal (BMI >=18.5 and <25) | 171 (15.2%) | 13 (11.1%) | 184 (14.8%) |  |
| 3: Overweight (BMI >=25 and <30) | 388 (34.6%) | 30 (25.6%) | 418 (33.7%) |  |
| 4: Obese (BMI >=30 and <40) | 479 (42.7%) | 60 (51.3%) | 539 (43.5%) |  |
| 5: Morbidly obese (BMI >=40) | 77 (6.9%) | 14 (12.0%) | 91 (7.3%) |  |
| Missing | 1 (0.1%) | 0 (0%) | 1 (0.1%) |  |
| **Body mass index (BMI continuous) (m/kg<sup>2</sup>)** |  |  |  |  |
| Mean (SD) | 30.6 (6.18) | 32.6 (7.45) | 30.8 (6.33) | 0.007 |
| Median [Min, Max] | 29.9 [15.1, 63.5] | 31.7 [19.9, 62.8] | 30.0 [15.1, 63.5] |  |
| Missing | 1 (0.1%) | 0 (0%) | 1 (0.1%) |  |
| **Waist Circumference (cm)** |  |  |  |  |
| Mean (SD) | 101 (14.7) | 106 (16.2) | 102 (14.9) | 0.002 |
| Median [Min, Max] | 100 [57.0, 178] | 104 [71.8, 166] | 101 [57.0, 178] |  |
| **Carotid Intima-Media Thickness (CiMT)** |  |  |  |  |
| Mean (SD) | 0.663 (0.167) | 0.727 (0.147) | 0.667 (0.167) |  |
| Median [Min, Max] | 0.634 [0.416, 2.23] | 0.672 [0.554, 1.04] | 0.638 [0.416, 2.23] |  |
| Missing | 865 (77.0%) | 99 (84.6%) | 964 (77.7%) |  |
| **Left Ventricular Internal Dimension at End-Diastole (LVIDd)** |  |  |  |  |
| Mean (SD) | 4.45 (0.446) | 4.62 (0.756) | 4.46 (0.472) | 0.48 |
| Median [Min, Max] | 4.40 [3.30, 5.60] | 4.70 [3.60, 6.10] | 4.40 [3.30, 6.10] |  |
| Missing | 970 (86.4%) | 106 (90.6%) | 1076 (86.8%) |  |
| **Left Ventricular Internal Dimension at End-Systole (LVIDs)** |  |  |  |  |
| Mean (SD) | 2.80 (0.344) | 2.95 (0.658) | 2.81 (0.372) | 0.49 |
| Median [Min, Max] | 2.80 [2.00, 3.80] | 2.80 [2.30, 4.40] | 2.80 [2.00, 4.40] |  |
| Missing | 970 (86.4%) | 106 (90.6%) | 1076 (86.8%) |  |
| **Left Ventricle Ejection Fraction** |  |  |  |  |
| Mean (SD) | 64.5 (4.01) | 65.9 (5.51) | 64.6 (4.12) | 0.43 |
| Median [Min, Max] | 64.3 [47.0, 74.3] | 63.8 [57.8, 73.0] | 64.0 [47.0, 74.3] |  |
| Missing | 977 (87.0%) | 106 (90.6%) | 1083 (87.3%) |  |
| **Left ventricle mass (2d_method)** |  |  |  |  |
| Mean (SD) | 135 (33.0) | 150 (30.4) | 136 (32.9) | 0.14 |
| Median [Min, Max] | 130 [68.9, 254] | 137 [111, 217] | 132 [68.9, 254] |  |
| Missing | 970 (86.4%) | 106 (90.6%) | 1076 (86.8%) |  |
| **Left Ankle-Brachial Index (ABI) (Dorsalis Pedis)** |  |  |  |  |
| Mean (SD) | 1.16 (0.142) | 1.11 (0.165) | 1.16 (0.143) | 0.47 |
| Median [Min, Max] | 1.18 [0.430, 1.47] | 1.14 [0.780, 1.33] | 1.17 [0.430, 1.47] |  |
| Missing | 1016 (90.5%) | 109 (93.2%) | 1125 (90.7%) |  |
| **Left Ankle-Brachial Index (ABI) (Posterior Tibial)** |  |  |  |  |
| Mean (SD) | 1.17 (0.183) | 1.11 (0.136) | 1.16 (0.180) | 0.33 |
| Median [Min, Max] | 1.16 [0.850, 2.46] | 1.13 [0.880, 1.31] | 1.16 [0.850, 2.46] |  |
| Missing | 1014 (90.3%) | 109 (93.2%) | 1123 (90.6%) |  |
| **Left Toe-Brachial Index (TBI)** |  |  |  |  |
| Mean (SD) | 0.968 (0.149) | 0.895 (0.120) | 0.963 (0.148) | 0.14 |
| Median [Min, Max] | 0.960 [0.670, 1.31] | 0.935 [0.680, | 0.955 [0.670, |  |
|  |  | 1.06] | 1.31] |  |
| Missing | 1015 (90.4%) | 109 (93.2%) | 1124 (90.6%) |  |
| **Right Ankle-Brachial Index (ABI) (Dorsalis Pedis)** |  |  |  |  |
| Mean (SD) | 1.16 (0.109) | 1.07 (0.153) | 1.15 (0.114) | 0.17 |
| Median [Min, Max] | 1.15 [0.890, 1.62] | 1.13 [0.730, 1.20] | 1.15 [0.730, 1.62] |  |
| Missing | 1012 (90.1%) | 109 (93.2%) | 1121 (90.4%) |  |
| **Right Ankle-Brachial Index (ABI) (posterior tibial)** |  |  |  |  |
| Mean (SD) | 1.14 (0.114) | 1.07 (0.187) | 1.14 (0.120) | 0.31 |
| Median [Min, Max] | 1.16 [0.740, 1.43] | 1.14 [0.640, 1.23] | 1.16 [0.640, 1.43] |  |
| Missing | 1015 (90.4%) | 109 (93.2%) | 1124 (90.6%) |  |
| **Right Toe-Brachial Index (TBI)** |  |  |  |  |
| Mean (SD) | 0.988 (0.273) | 1.10 (0.446) | 0.995 (0.287) | 0.52 |
| Median [Min, Max] | 0.930 [0.480, 1.97] | 1.07 [0.550, 2.04] | 0.935 [0.480, 2.04] |  |
| Missing | 1013 (90.2%) | 109 (93.2%) | 1122 (90.5%) |  |
| **Low Density Lipoprotein (LDL-c)** |  |  |  |  |
| Mean (SD) | 111 (33.9) | 103 (36.5) | 110 (34.2) | 0.03 |
| Median [Min, Max] | 108 [23.0, 270] | 107 [10.0, 270] | 108 [10.0, 270] |  |
| Missing | 17 (1.5%) | 1 (0.9%) | 18 (1.5%) |  |
| **High Density Lipoprotein (HDL-c)** |  |  |  |  |
| Mean (SD) | 47.6 (12.8) | 45.5 (12.6) | 47.4 (12.8) | 0.09 |
| Median [Min, Max] | 46.0 [3.00, 111] | 46.0 [0, 87.0] | 46.0 [0, 111] |  |
| Missing | 8 (0.7%) | 1 (0.9%) | 9 (0.7%) |  |
| **Total Cholesterol** |  |  |  |  |
| Mean (SD) | 187 (41.4) | 181 (38.8) | 187 (41.2) | 0.08 |
| Median [Min, Max] | 185 [52.0, 390] | 176 [106, 274] | 185 [52.0, 390] |  |
| Missing | 9 (0.8%) | 1 (0.9%) | 10 (0.8%) |  |
| **Diastolic Blood Pressure (mmHg)** |  |  |  |  |
| Mean (SD) | 71.5 (9.45) | 71.1 (9.25) | 71.4 (9.43) | 0.73 |
| Median [Min, Max] | 71.0 [40.0, 113] | 71.0 [46.0, 99.0] | 71.0 [40.0, 113] |  |
| Missing | 2 (0.2%) | 0 (0%) | 2 (0.2%) |  |
| **Systolic Blood Pressure (mmHg)** |  |  |  |  |
| Mean (SD) | 117 (16.7) | 122 (16.6) | 117 (16.7) | 0.001 |
| Median [Min, Max] | 114 [80.0, 211] | 119 [86.0, 180] | 114 [80.0, 211] |  |
| Missing | 2 (0.2%) | 0 (0%) | 2 (0.2%) |  |
| **C-Reactive Protein (CRP)** |  |  |  |  |
| Mean (SD) | 9.07 (16.8) | 7.27 (9.52) | 8.87 (16.2) | 0.12 |
| Median [Min, Max] | 3.80 [0.0400, 115] | 5.30 [0.500, 77.7] | 3.89 [0.0400, 115] |  |
| Missing | 335 (29.8%) | 21 (17.9%) | 356 (28.7%) |  |
| **Interleukin 6 (IL_6)** |  |  |  |  |
| Mean (SD) | 8.62 (116) | 5.24 (10.6) | 8.28 (110) | 0.46 |
| Median [Min, Max] | 2.22 [0.0275, 3040] | 2.39 [0.478, 77.5] | 2.28 [0.0275, 3040] |  |
| Missing | 430 (38.3%) | 39 (33.3%) | 469 (37.8%) |  |
| **Interleukin 1 Beta (IL_1Beta)** |  |  |  |  |
| Mean (SD) | 1.09 (3.52) | 1.14 (2.12) | 1.09 (3.40) | 0.87 |
| Median [Min, Max] | 0.640 [0.00217, 67.3] | 0.640 [0.0252, 13.7] | 0.640 [0.00217, 67.3] |  |
| Missing | 443 (39.4%) | 39 (33.3%) | 482 (38.9%) |  |
| **Interleukin 8 (IL8)** |  |  |  |  |
| Mean (SD) | 6.70 (7.39) | 5.62 (3.89) | 6.59 (7.12) | 0.04 |
| Median [Min, Max] | 4.69 [0.0458, 81.7] | 4.43 [0.640, 25.0] | 4.66 [0.0458, 81.7] |  |
| Missing | 429 (38.2%) | 39 (33.3%) | 468 (37.7%) |  |
| **rs429358 (chr19:44908684)** |  |  |  |  |
| Major allele T (TT) | 873 (77.7%) | 94 (80.3%) | 967 (78.0%) | 0.69 |
| Minor allele C (CT or CC) | 206 (18.3%) | 20 (17.1%) | 226 (18.2%) |  |
| Missing | 44 (3.9%) | 3 (2.6%) | 47 (3.8%) |  |
| **rs7412 (chr19:44908822)** |  |  |  |  |
| Major allele C (CC) | 1002 (89.2%) | 110 (94.0%) | 1112 (89.7%) | 0.14 |
| Minor allele T<br>(CT or TT) | 77 (6.9%) | 4 (3.4%) | 81 (6.5%) |  |
| Missing | 44 (3.9%) | 3 (2.6%) | 47 (3.8%) |  |
| **APOE haplotype ε4** |  |  |  |  |
| ε4 absent | 878 (78.2%) | 94 (80.3%) | 972 (78.4%) | 0.88 |
| ε4 present | 201 (17.9%) | 20 (17.1%) | 221 (17.8%) |  |
| Missing | 44 (3.9%) | 3 (2.6%) | 47 (3.8%) |  |
| **APOE haplotypes** |  |  |  |  |
| ε4ε4 | 14 (1.2%) | 4 (3.4%) | 18 (1.5%) | 0.26 |
| ε3ε4 | 187 (16.7%) | 16 (13.7%) | 203 (16.4%) |  |
| ε2ε4 | 5 (0.4%) | 0 (0%) | 5 (0.4%) |  |
| ε3ε3 | 801 (71.3%) | 90 (76.9%) | 891 (71.9%) |  |
| ε2ε3 | 71 (6.3%) | 4 (3.4%) | 75 (6.0%) |  |
| ε2ε2 | 1 (0.1%) | 0 (0%) | 1 (0.1%) |  |
| Missing | 44 (3.9%) | 3 (2.6%) | 47 (3.8%) |  |

### DNA Genotyping and Imputation

DNA genotyping and imputation procedures for the CCHC cohort were previously described [10]. Briefly, Illumina chip (Multi-Ethnic Genotyping Array – MEGA) was used to provide genotypes specific to Hispanic population. After quality control, genotypes were imputed using the TOPMed reference panel and the Michigan Imputation Server [10]. For the *APOE* haplotype analyses, we used two imputed SNPs (rs429358 and rs7412) in the *APOE* region (imputation R^2^>0.95) to estimate the *APOE* haplotype in our cohort using PLINK and R [11].

### Results

The total number of subjects 18 years and older with complete follow-up data on MMSE at baseline of the study was 3,761. A total of 1,240 out of 3,761 subjects completed the 5-year interview and were available for the current analysis. Baseline study cohort characteristics are shown in Table 1. A total of 117 out of 1,240 subjects showed cognitive impairment (MMSE<24) at study baseline. The majority of the subjects involved from this cohort were less than 65 years old (89.9%) and the male-to-female ratio in this cohort was about 2 to 1. Table 1. Subjects with cognitive impairment were more likely to be older in age, have lower education levels, have a history of myocardial infarction or stroke. Table 1. Having heterozygous (*APOE4E3*) or homozygous (*APOE4E4*) *APOE* haplotypes did not significantly increase the risk for cognitive impairment in our cohort. Table 1. *APOE* haplotypes distributions are shown in Table 1.

The mixed logistic regression models’ estimates for crude and adjusted odds ratios are shown in Table 2. We adjusted our modes for age (categories:18-49, 50-64, ≥65), education level (categories: ≥13, 9-12, 5-8, ≤4), history of incident stroke, and *APOE4* haplotype status. Table 2. Diabetes mellitus significantly increased the odds for cognitive impairment by 2 times (OR:2.11, 95%CI (1.26-3.52)) when compared to those without Diabetes and after adjusting for age, education, history of stroke, and *APOE4* haplotype status. Table 2. Lower LDL-c levels, higher HDL-c levels, and higher systolic blood pressure decreased the odds for cognitive impairment; while higher diastolic blood pressure, higher BMI, and metabolic syndrome increased the odds for cognitive impairment. However, besides diabetes, none of those cardiometabolic risk factors remained significant after adjusting for multiple testing. Table 2. We tested for a dose-response relationship between glycosylated Hemoglobin A1c (HBA1c) and cognitive impairment using unadjusted and adjusted mixed logistic regression modeling. Table 3. Our dose-response analyses showed a significant dose-response association between increasing levels of HBA1c and having cognitive impairment. Table 3.

**Table 2.** Associations from several cardiometabolic indicators with cognitive impairment from crude and adjusted mixed logistic regression models. False Discovery Rate (FDR) was used to adjust for multiple testing. All models were adjusted for age, level of education, prior stroke, and presence of *APOE* ɛ4 haplotype.

| **Risk Factor** | **Unadjusted**<br>**Odds Ratio (95%CI)** | **Adjusted**<br>**Odds Ratio (95%CI)** | **FDR\*** |
| --- | --- | --- | --- |
| **Diabetes** | 6.46<br>(3.69-11.29)<br>$5.83 \times 10^{-11}$ | 2.11<br>(1.26- 3.52)<br>0.004 | 0.03 |
| **Low Density**<br>**Lipoprotein (LDL-c)** | 0.46<br>(0.25-0.82)<br>0.009 | 0.62<br>(0.35-1.09)<br>0.10 | 0.26 |
| **High Density**<br>**Lipoprotein (HDL-c)** | 1.64<br>(1.03- 2.62)<br>0.04 | 1.34<br>(0.85-2.10)<br>0.21 | 0.36 |
| **Total Cholesterol** | 0.79<br>(0.48-1.31)<br>0.36 | 0.83<br>(0.51-1.36)<br>0.47 | 0.52 |
| **Systolic Blood**<br>**Pressure** | 1.72<br>(1.01-2.93)<br>0.05 | 0.85<br>(0.50-1.44)<br>0.55 | 0.55 |
| **Diastolic Blood**<br>**Pressure** | 1.07<br>(0.58-1.99)<br>0.82 | 1.36<br>(0.75-2.48)<br>0.31 | 0.41 |
| **Body Mass Index**<br>**(BMI)** | 1.46<br>(0.70-3.01)<br>0.31 | 1.34<br>(0.67-2.69)<br>0.41 | 0.47 |
| **Waist**<br>**Circumference** | 1.83<br>(1.11-3.00)<br>0.02 | 1.33<br>(0.84-2.12)<br>0.22 | 0.36 |
| **Metabolic Syndrome** | 2.90<br>(1.81-4.63)<br>$8.66 \times 10^{-6}$ | 1.64<br>(1.04-2.57)<br>0.03 | 0.13 |

FDR: false discovery rate (p value <0.5 as threshold for significance)

**Table 3.** Shows the dose-response associations using unadjusted and adjusted mixed logistic regression modeling.

| **HBA1c** | **Unadjusted Odds Ratio (95%CI)** | **p value** | **Adjusted Odds Ratio (95%CI)** | **p value** |
| --- | --- | --- | --- | --- |
| **<5** | 1 | ... | 1 | ... |
| **≥5-<6.5** | 1.09<br>(0.62-1.91) | 0.77 | 1.13<br>(0.64-1.99) | 0.66 |
| **≥6.5-<8** | 6.08<br>(2.74-13.50) | $8.96 \times 10^{-6}$ | 2.32<br>(1.06-5.06) | 0.03* |
| **≥8** | 3.47<br>(1.56-7.72) | 0.002 | 1.29<br>(0.59-2.82) | 0.52 |

## Discussion

We showed that the cognitive impairment process in the majority of our Mexican American H/L subjects develops below the age of 65, and is associated with metabolic risk factors, particularly diabetes. This important observation opens possibilities for establishing early biomarkers and preventive approaches to lower the burden of cognitive impairment in H/L populations, mainly Mexican Americans. The current analysis was a longitudinal study to investigate the associations of several cardiometabolic risk factors with cognitive impairment in a relatively young Hispanic population of mainly Mexican Americans living in the Rio Grande Valley. We adjusted our analyses for age, level of education, the presence of a self-reported incident stroke, and *APOE* haplotype status. Our adjusted models and, after correction for multiple testing (FDR p value <0.05), showed that diabetes was positively associated with incident cognitive impairment in our population. Low-Density lipoproteins (LDL-c), High-density lipoproteins (HDL-c), total cholesterol (TC), metabolic syndrome, and both systolic and diastolic blood pressures were not associated with cognitive impairment in our adjusted analyses.

The diabetes association with cognitive impairment is well-established in prior research studies of H/L and non-H/L populations. Our results showed that participants with diabetes had two times the odds of incident cognitive impairment compared to those without diabetes after adjusting for age, education level, history of incident stroke, and *APOE* haplotype (OR: 2.11, 95%CI: 1.16-2.63). Prior research by Gonzalez et al. reported a 74% increase in the odds of having prevalent diabetes among those with cognitive impairment in middle-aged and elderly Hispanics from the Study of Latinos cohort (odds ratio 1.74 [95% CI 1.34; 2.26]; *P* < 0.001) [7], and a weaker association with incident diabetes and cognition [7]. Similarly, prior research on diabetes and cognition in older H/L (≥70 years of age) showed increased risk for cognitive impairment among those with diabetes when compared to non-diabetics [12]. While prior research was focused on older Hispanics over the age of 65 years, our younger Hispanic population (89.9% below that age of 65) showed that over 40% of those with impaired cognitive function were between the age of 50 to 64 years-old. Table 1. Furthermore, we were able to show a significant (p value<0.05) dose-response relationship between increasing levels of HBA1c and having cognitive impairment, a novel finding in Mexican American H/L [13]. Table 3.

Genetic studies on *APOE* and diabetes genes and their associations with cognitive impairment and Alzheimer disease have been inconclusive among Hispanics. A study from the Million Veteran Program showed no associations between genetic risk score for diabetes genes and dementia among Hispanics, regardless of the *APOE* allele status. [6]. *APOE* haplotypes showed the weakest associations with Alzheimer’s disease when compared to *APOE* haplotypes associations with Alzheimer’s disease from Asians, Non-H/L Whites, and Non-Hispanic Black populations [3, 14]. This attenuated association between *APOE*ɛ4 alleles and dementia could be attributed to the lower prevalence of *APOE* alleles in H/L populations [5]. Our findings from analyzing the *APOE* haplotype in our cohort showed no association between having an *APOE*ɛ4 haplotype and risk for cognitive impairment and were in line with previous analyses in Hispanic/Latino populations [15–17]. Table 1. This may indicate that the increased risk for cognitive impairment in diabetic Hispanics is mainly driven by the cardiometabolic effects of diabetes on brain vasculature [14].

Our analyses investigated the associations of several lipid profiles (e.g., LDL-c, HDL-c, and Total Cholesterol) with cognitive impairment. LDL-c mean and median were significantly lower among those with cognitive impairment compared to those with normal cognition in our cohort. Although statistically insignificant, our adjusted models showed that a higher LDL-c to have protective effect on cognition (OR:0.62, 95%CI: 0.35-1.09). Prior reports on lipid profiles and cognition in H/L were inconsistent. Similar to our results, Reitz at el., reported sex and age adjusted associations between LDL-c level and incident cognitive impairment that were protective for incident cognitive impairment among Hispanics 65 years and older [18]. However, this protective LDL association with cognitive impairment was attenuated after adjusting for education, AOPE4, age, sex, and vascular risk factors [18]. Reitz at el., concluded that there were no associations between lipids (LDL-c, HDL-c, TC) and incident cognitive impairment among those 65 and older. A more recent cross-sectional study from the Study of Latinos cohort showed that higher total cholesterol and LDL-c levels were associated with improved learning and verbal cognition [19], an unusual finding that is in line with our report on the association between LDL-c and cognition in our longitudinal analyses. Furthermore, this study showed an effect modification of Hispanic background in the association of LDL-c and cognition suggesting that different H/L backgrounds have different effects on this association [19]. Adjusting for antihyperlipidemic medications did not change the association between LDL-c and cognition [19]. Similar to our findings on the association between HDL-c and cognition from our cohort, Lamar et al., showed that HDL-c to be insignificantly associated with cognitive test performance in learning, memory, and verbal fluency domains [19].

Hypertension is an important risk factor for vascular health and vascular cognitive impairment. Few studies had examined the associations of having hypertension on cognitive impairment among Hispanics. Márquez et al., reported that having persistent hypertension (i.e., prevalent hypertension) was significantly associated with cognitive decline in the Study of Latinos Hispanic cohort; however, incident hypertension in the same cohort was not associated with cognitive impairment [20]. Although our study did not differentiate between persistent and incident hypertension, our adjusted model did not show a significant association of either systolic or diastolic blood pressure measurements with incident mild cognitive decline. This lack of association between blood pressure readings and incident mild cognitive impairment in our cohort may be explained by the fact that our observations for blood pressure were applied at one time point per visit with no continuous or serial blood pressure readings in-between the visits. Furthermore, Marquez et al., included a mixture of Hispanic/Latino populations in their analyses with only 34.4% were Mexican Americans, and did not stratify their hypertension associations with cognitive impairment by country of origin [20]. Different H/L populations might contribute to the differences in the reported associations between hypertension and cognitive impairment from our cohort and those reported by Marquez et al.

### Strengths and Limitations

Our study has several strengths such as the longitudinal study design and the random recruitment from the population that was not based on the presence of cognitive impairment. Several limitations in our analyses included the use of the MMSE which is known for its low sensitivity in detecting cognitive impairment [21], may resulted in missed opportunities to identify cases of cognitive impairment. We are currently improving our cognitive testing screening procedures in the CCHC cohort through implementing the Montreal Cognitive Assessment (MOCA) test which is proven for its higher sensitivity in detecting mild cognitive impairment over MMSE [21]. Although we used high quality imputed SNPs to estimate our *APOE* haplotypes; the attenuated *APOE4* associations in our cohort may be related to the use of imputed SNPs instead of genotyped or sequenced SNPs to estimate *APOE* alleles frequencies. Missing values from several important cardiometabolic indicators such as echocardiography, carotid intimal thickness, and inflammatory markers measurements might result in missing opportunities in identifying associations with cognitive impairment. The relatively small sample size may also contribute to the low power of detecting significant associations.

### Conclusions

Diabetes showed significant association with MCI in young and middle-aged Hispanics in our longitudinal study. This, in addition to the lower impact of *APOE* haplotypes on Hispanic populations, makes it possible that main drivers for cognitive impairment in this population most likely are driven by vascular pathologies related to diabetes and its related cardiometabolic outcomes. Future studies should examine the link between vessel health of the cerebrovasculature and cognition in Hispanic patients less than 65 years old

## Data Availability

Data available upon request from Dr. Joseph McCormick and Dr. Susan Fisher-Hoch.

## References

1. Wang, X., et al., Predictive models of Alzheimer’s disease dementia risk in older adults with mild cognitive impairment: a systematic review and critical appraisal. BMC Geriatr, 2024. 24(1): p. 531.

2. Rundek, T., et al., Vascular Cognitive Impairment (VCI). Neurotherapeutics, 2022. 19(1): p. 68–88.

3. Belloy, M.E., et al., APOE Genotype and Alzheimer Disease Risk Across Age, Sex, and Population Ancestry. JAMA Neurol, 2023. 80(12): p. 1284–1294.

4. Farrer, L.A., et al., Effects of age, sex, and ethnicity on the association between apolipoprotein E genotype and Alzheimer disease. A meta-analysis. APOE and Alzheimer Disease Meta Analysis Consortium. Jama, 1997. 278(16): p. 1349–56.

5. O’Bryant, S.E., et al., Characterization of Mild Cognitive Impairment and Dementia among Community-Dwelling Mexican Americans and Non-Hispanic Whites. J Alzheimers Dis, 2022. 90(2): p. 905–915.

6. Litkowski, E.M., et al., A Diabetes Genetic Risk Score Is Associated With All-Cause Dementia and Clinically Diagnosed Vascular Dementia in the Million Veteran Program. Diabetes Care, 2022. 45(11): p. 2544–2552.

7. González, H.M., et al., Diabetes, Cognitive Decline, and Mild Cognitive Impairment Among Diverse Hispanics/Latinos: Study of Latinos-Investigation of Neurocognitive Aging Results (HCHS/SOL). Diabetes Care, 2020. 43(5): p. 1111–1117.

8. Fisher-Hoch, S.P., et al., Socioeconomic status and prevalence of obesity and diabetes in a Mexican American community, Cameron County, Texas, 2004-2007. Prev Chronic Dis, 2010. 7(3): p. A53.

9. Bukhbinder, A.S., et al., Population-Based Mini-Mental State Examination Norms in Adults of Mexican Heritage in the Cameron County Hispanic Cohort. J Alzheimers Dis, 2023. 92(4): p. 1323–1339.

10. Sabotta, C.M., et al., Genetic variants associated with circulating liver injury markers in Mexican Americans, a population at risk for non-alcoholic fatty liver disease. Front Genet, 2022. 13: p. 995488.

11. Lumsden, A.L., et al., Apolipoprotein E (APOE) genotype-associated disease risks: a phenome-wide, registry-based, case-control study utilising the UK Biobank. EBioMedicine, 2020. 59: p. 102954.

12. Luchsinger, J.A., et al., Relation of diabetes to mild cognitive impairment. Arch Neurol, 2007. 64(4): p. 570–5.

13. González, H.M., et al., Glycemic Control, Cognitive Aging, and Impairment Among Diverse Hispanic/Latino Individuals: Study of Latinos-Investigation of Neurocognitive Aging (Hispanic Community Health Study/Study of Latinos). Diabetes Care, 2024. 47(7): p. 1152–1161.

14. Haan, M.N., et al., Prevalence of dementia in older latinos: the influence of type 2 diabetes mellitus, stroke and genetic factors. J Am Geriatr Soc, 2003. 51(2): p. 169–77.

15. Huggins, L.K.L., et al., Meta-Analysis of Variations in Association between APOE ɛ4 and Alzheimer’s Disease and Related Dementias Across Hispanic Regions of Origin. J Alzheimers Dis, 2023. 93(3): p. 1095–1109.

16. Campos, M., S.D. Edland, and G.M. Peavy, Exploratory study of apolipoprotein E ε4 genotype and risk of Alzheimer’s disease in Mexican Hispanics. J Am Geriatr Soc, 2013. 61(6): p. 1038–1040.

17. O’Bryant, S.E., et al., Risk factors for mild cognitive impairment among Mexican Americans. Alzheimers Dement, 2013. 9(6): p. 622–631.e1.

18. Reitz, C., et al., Plasma lipid levels in the elderly are not associated with the risk of mild cognitive impairment. Dement Geriatr Cogn Disord, 2008. 25(3): p. 232–7.

19. Lamar, M., et al., Associations of Lipid Levels and Cognition: Findings from the Hispanic Community Health Study/Study of Latinos. J Int Neuropsychol Soc, 2020. 26(3): p. 251–262.

20. Márquez, F., et al., Hypertension, Cognitive Decline, and Mild Cognitive Impairment Among Diverse Hispanics/Latinos: Study of Latinos-Investigation of Neurocognitive Aging Results (SOL-INCA). J Alzheimers Dis, 2024. 97(3): p. 1449–1461.

21. Pendlebury, S.T., et al., Differences in cognitive profile between TIA, stroke and elderly memory research subjects: a comparison of the MMSE and MoCA. Cerebrovasc Dis, 2012. 34(1): p. 48–54.

